# When Is Amyloid Really Cleared? Substantial Discordance Between Centiloid Quantitation and Visual Reads of Amyloid PET after Amyloid-Targeting Therapy

**DOI:** 10.64898/2026.10.03.26364642

**Authors:** Renaud La Joie, Marlene Lin, Yingbing Wang, David N. Soleimani-Meigooni, Tara Ellingson, Hong Nguyen, Ganna Blazhenets, Corrina S. Fonseca, Kambiz Nael, Salil Soman, Pierrick Bourgeat, Jake P. Levy, Gina Rhee, Julio C. Rojas, Melanie Stephens, Nhat Bui, Peter A. Ljubenkov, Lawren VandeVrede, Gil D. Rabinovici

## Abstract

Amyloid-targeting therapies (ATTs) substantially reduce amyloid PET signal and slow clinical decline in early symptomatic Alzheimer disease. As ATTs enter clinical practice, PET may assess target engagement and guide treatment management, but there is no consensus on how to measure treatment-related amyloid clearance (TRAC). We compared Centiloid quantitation across pipelines and visual reads for interpreting pre- and post-ATT amyloid PET.

**Methods:** We analyzed 18F-florbetapir or 18F-florbetaben PET from 101 patients treated with lecanemab or donanemab: 88 pre-ATT and 54 post-ATT scans, including 41 paired scans. Centiloids were derived using FDA-cleared MIMneuro and two research pipelines (rPOP, CapAIBL) and compared using intraclass correlation coefficients (ICC). Clearance rates were estimated using linear mixed-effects models. Full TRAC was defined on post-ATT scans quantitatively (<11 Centiloids) or based on visual interpretation by 3 expert readers.

**Results:** Treated patients were 74±7 years old; 57% were female, 59% were apolipoprotein E ε4 carriers, and 78% had mild cognitive impairment (22% mild dementia). Centiloid agreement between pipelines was equally high pre- and post-ATT (both ICCs=0.90). Among paired scans (mean treatment duration, 16.7±4.6 months), clearance estimates were similar across pipelines and greater with donanemab (n=15 patients; between -62 and -60 Centiloids/year depending on the pipeline) than lecanemab (n=26; -38 to -36 Centiloids/year; all P<0.005). Among 54 post-ATT scans, between 44% and 57% were <11 Centiloids depending on pipeline, with 20 (37%) meeting this criterion across all 3 pipelines. In contrast, only 11%–19% were visually negative across readers, with 4 (7%) unanimously read as negative. Of the 20 post-ATT scans <11 Centiloids across all pipelines, 13 (65%) were considered positive by all readers, commonly with residual occipital binding.

**Conclusion:** Centiloid quantitation was robust across pipelines before and after ATT initiation, yielding clearance rates consistent with clinical trials. However, quantitative and visual assessments were markedly discordant after ATT, with an unusually high proportion of scans <11 Centiloids remaining visually positive, warranting cautious interpretation of post-ATT Centiloid values. Larger studies should assess the clinical relevance of these findings and further develop robust approaches to establish full TRAC.

## Introduction

Amyloid-targeting therapies (ATTs) have transformed the clinical landscape of Alzheimer disease (AD), by demonstrating a modest slowing of clinical decline in patients at the early clinical stages of AD(*1*,*2*). Amyloid PET has supported ATT development by both confirming pre-ATT amyloid positivity and demonstrating treatment-related amyloid clearance (TRAC)(*3*). In randomized controlled trials of lecanemab and donanemab, the two ATTs currently used in clinical practice, amyloid PET became negative (“full TRAC”) in approximately two-thirds of participants after 18 months of treatment, based on trial-specific criteria(*3*). In the donanemab trials, amyloid PET were repeated every 6 months to adapt treatment duration: patients initially randomized to donanemab were switched to placebo when the trial’s criteria for full TRAC was met (*1*,*4*). As ATTs enter clinical practice, repeated amyloid PET may provide evidence of pharmacodynamic changes and ultimately inform treatment duration or discontinuation, but no consensus exists on how to operationalize TRAC in clinical practice(*3*).

In clinical settings, amyloid PET has traditionally been interpreted visually, resulting in a binary positive or negative classification. Recent changes to FDA labels and updates to the appropriate use criteria(*5*) have supported the use of PET quantitation as an adjunct to visual interpretation. The most widely adopted approach is to quantify amyloid PET signal in Centiloids, a standardized scale that facilitates comparisons across radiotracers and analysis methods(*6*). Quantitation may be particularly informative after ATTs to characterize TRAC beyond a binary positive/negative outcome.

Several tools are now available to calculate Centiloid values, including clinical software cleared by regulatory agencies and research pipelines that have demonstrated high agreement (*7–9*). Similarly, visual interpretation and Centiloid quantitation are highly concordant (kappa=0.86 in a meta-analysis of 36 studies/35,045 participants(*10*)). However, these validation studies have been conducted in ATT-naive individuals, and it remains unclear whether the same level of agreement is maintained after ATT, when marked reductions in cortical signal and potential changes in regional tracer distribution, brain volume, and perfusion, that may alter PET image characteristics.

As longitudinal amyloid PET is increasingly used to assess treatment response, it is important to test empirically whether different interpretation approaches remain reliable after treatment, to inform clinically applicable strategies for assessing TRAC. In this study, we evaluated amyloid PET scans acquired before and after ATT initiation to assess the reliability and agreement of complementary approaches for characterizing TRAC. Specifically, we compared Centiloid quantitation across 3 image analysis pipelines (1 FDA-cleared software and 2 widely used research pipelines) and examined their agreement with expert visual reads.

## Methods

### Participants

The participants enrolled in this study were approved for ATT by the University of California, San Francisco (UCSF) Alzheimer infusion clinic adjudication committee based on appropriate use recommendations(*11*,*12*) and clinical experience. Patients underwent comprehensive neurological evaluation including review of prior medical history, Apolipoprotein E (*APOE*) genotype, 3T-MRI, complete neurological exam, and assessment of clinical stage. Participants were treated with intravenous lecanemab every two weeks (starting in October 2023) or donanemab every four weeks (starting in November 2024), per FDA-approved label.

All participants provided written consent, before ATT initiation, for their clinical data to be used for research. The study was approved by the UCSF Institutional Review Board.

### PET acquisition and processing

PET scans were performed with 18F-florbetapir (n=136 scans; 38.9 ± 8.2 min post injection) or 18F-florbetaben (n=6 scans, all pre-ATT; 91.2 ± 1.9 min post injection) and acquired on Siemens (n=73 scans), Philips (n=58), GE HealthCare (n=10) or United Imaging Healthcare (n=1) PET/CT scanners.

PET scans were quantified in Centiloids using 3 independent pipelines. First, an FDA-cleared software, MIMneuro (v7.3.5, Amyloid Centiloid Analysis Workflow, MIM Software Inc., Cleveland, OH, USA), was implemented by the UCSF Nuclear Medicine team and included in the interpretation of clinical amyloid PET. For research pipelines, DICOM files were converted to NIfTI; multiframe acquisitions were realigned, averaged, and processed using rPOP(*13*) and CapAIBL(*14*). All 3 tools are PET-only pipelines that do not require structural imaging.

### Operationalization of TRAC classification

Post-ATT scans were classified in two complementary ways to identify “full TRAC”, i.e., an amyloid-negative post-ATT scan. First, to define full TRAC quantitatively, we applied a stringent threshold of <11 Centiloids. This value was i) used in the TRAILBLAZER-ALZ2 trial to stop active donanemab treatment (*1*) and ii) identified in a meta-analysis of 53 studies (49,227 participants) as a high certainty negativity threshold (*10*). This value is also very close to the 12-Centiloid threshold identified in a PET-to-autopsy study to detect neuritic plaques(*15*). Second, post-ATT scans (all 18F-florbetapir PET) were visually interpreted as positive or negative by 3 independent expert readers (RLJ, DNSM, YW) following the method described in the radiotracer prescribing information. Scans were considered positive if two or more cortical areas showed reduced or absent gray/white contrast, or if one or more areas showed gray matter signal clearly exceeding that of adjacent white matter. Readers were blinded to clinical information, Centiloid values, and structural imaging, but were aware that scans were acquired after ATT initiation.

In addition, we aimed to identify patients with no TRAC, i.e., patients whose post-ATT scan showed no meaningful Centiloid decline relative to the pre-ATT scan. No TRAC was defined as either increase in Centiloid or a decrease within the range of test-retest variability, which was operationalized as a decline smaller than 15 Centiloids based on previous analyses(*16*).

### Additional dataset for complementary analyses

To contextualize findings from the current study, we aimed to document the relationship between binary visual reads and Centiloid quantitation (using an 11 Centiloid threshold) in ATT-naive participants using the Imaging Dementia—Evidence for Amyloid Scanning (IDEAS) study (*17*). IDEAS enrolled cognitively impaired Medicare beneficiaries who met the 2013 appropriate use criteria for amyloid PET (*18*) between 2016 and 2018, i.e., prior to both ATT approval and use of Centiloid in clinical settings. Scans were acquired with one of 3 FDA-approved radiotracers at 343 PET facilities across the US and visually interpreted by imaging specialists (board-certified radiologist or nuclear medicine physician) at each site. We previously quantified 10,361 of these scans in Centiloids using rPOP(*19*). To maximize comparability with our post-ATT scans, we selected IDEAS scans that were i) acquired with 18F-florbetapir, ii) quantified <11 Centiloids, and iii) had a binary (positive or negative) visual read, yielding 2,265 scans.

### Statistical analyses

Statistical analyses were performed using R software (version 4.5.1, R Core Team, 2025). Agreement across pipelines was assessed using intraclass correlation coefficients (ICC) based on a two-way model, absolute agreement, and single-measure reliability (R package *irr*). Analyses were run separately for pre-ATT and post-ATT scans to evaluate whether the reliability of Centiloid estimation was affected by ATT.

In participants with both pre- and post-ATT PET, amyloid clearance rates were estimated using linear mixed-effects models of Centiloid values as a function of time since ATT initiation, with random intercepts (R package *lme4*). Subsequent models compared clearance rates between lecanemab- and donanemab-treated patients by adding a time*drug interaction. Analyses were run separately for each pipeline.

For analyses of post-ATT scans assessing the full TRAC status, agreement between the classifications (3 pipelines or 3 visual readers) was quantified using Fleiss’ kappa values, or Cohen’s kappa when reporting pairwise comparisons.

## Results

### Sample

Among 101 participants with mild cognitive impairment or mild dementia treated at UCSF (see Table 1 for demographic and clinical characteristics), 88 had pre-ATT PET, 54 post-ATT PET, and 41 had paired scans; 47 only had pre-ATT PET and 13 only had post-ATT PET.

**Table 1.** Characteristics of participants included in the study.

|  | Overall sample<br>(n=101) | Subgroup with pre-ATT scans (n=88) | Subgroup with post-ATT scans (n=54) |
| --- | --- | --- | --- |
| **Age, years** | 73.7 ± 6.7 | 73.9 ± 6.3 | 73.2 ± 7.1 |
| **Sex, n (%)** |  |  |  |
| Female | 58 (57%) | 50 (57%) | 32 (59%) |
| Male | 43 (43%) | 38 (43%) | 22 (41%) |
| **Education, years** | 17.1 ± 2.2 | 17.0 ± 2.3 | 17.4 ± 2.1 |
| **APOE ε4 status, n (%)** |  |  |  |
| Carrier | 60 (59%) | 53 (60%) | 28 (52%) |
| Non-carrier | 41 (41%) | 35 (40%) | 26 (48%) |
| **Diagnosis, n (%)** |  |  |  |
| MCI | 79 (78%) | 70 (80%) | 41 (76%) |
| Mild dementia | 22 (22%) | 18 (20%) | 13 (24%) |
| **CDR-SB** | 2.7 ± 1.4 | 2.7 ± 1.4 | 2.8 ± 1.4 |
| **MMSE** | 25.4 ± 3.1 | 25.4 ± 3.1 | 25.6 ± 2.7 |
| **Radiotracer** |  |  |  |
| 18F-florbetaben | - | 6 (7%) | 0 |
| 18F-florbetapir | - | 82 (93%) | 54 (100%) |
| **Treatment** |  |  |  |
| Lecanemab | - | - | 39 (72%) |
| Donanemab | - | - | 15 (28%) |
| **Treated duration, months** | - | - | 17.1 ± 4.6 |

All demographic and clinical characteristics describe baseline (pre-ATT) state.

### Centiloid quantitation

Agreement between Centiloid values across the 3 pipelines was high for both pre-ATT scans (n=88, ICC = 0.899, 95%CI [0.848 to 0.934], all pairwise ICCs ≥ 0.87, Figure 1A) and post-ATT scans (n=54, ICC= 0.902 [0.812 to 0.946], all pairwise ICCs ≥ 0.90, Figure 1B).

**Figure 1.**
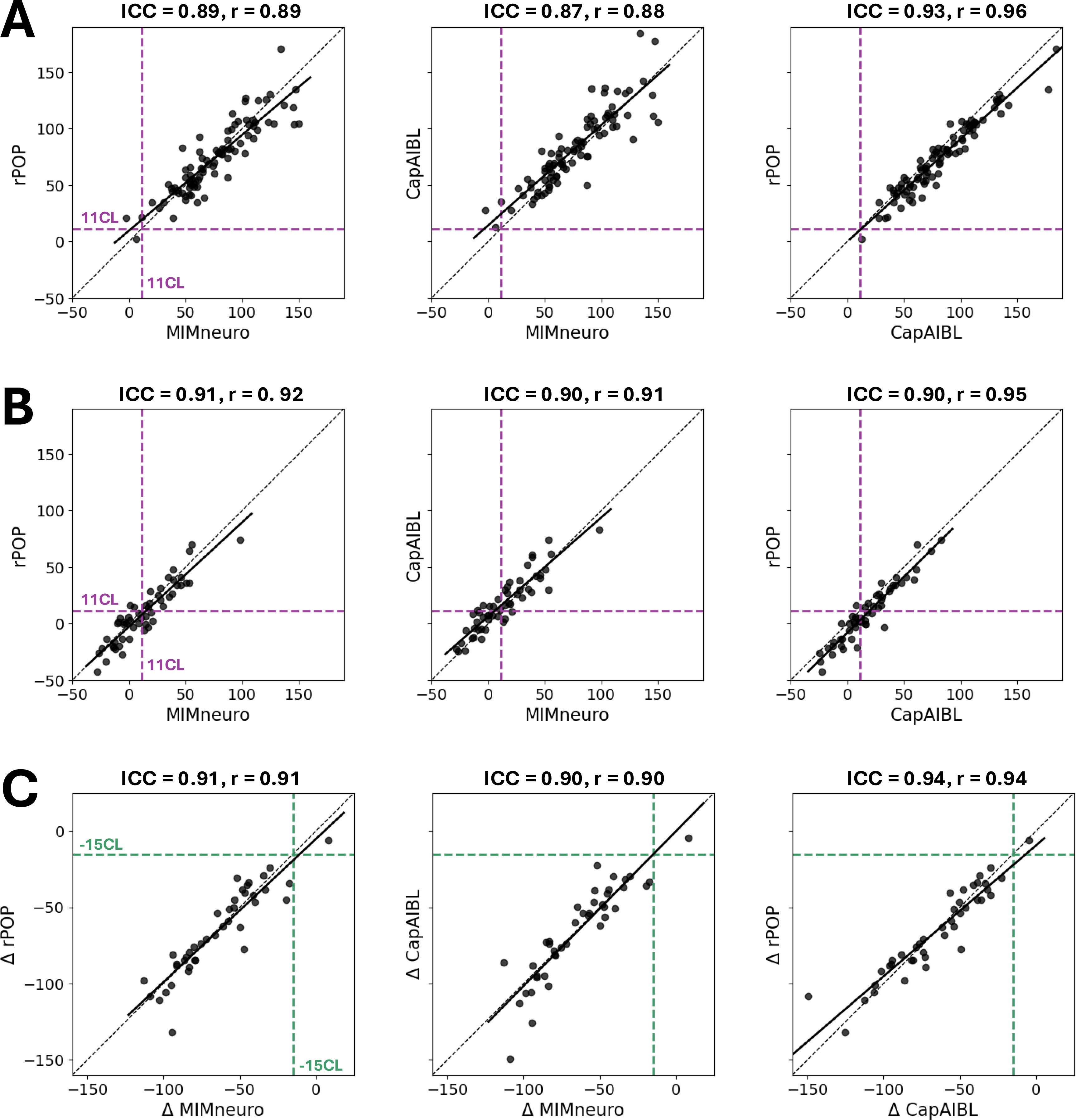
Agreement between Centiloid values across pipelines. A) Scans acquired before ATT initiation (n=88). B) Scans acquired after ATT initiation (n=54). C) Change in Centiloids calculated for individuals with paired scans (n=41). Black dashed lines: identity; Black solid lines: regression lines; Purple dashed lines (A-B): 11 Centiloid threshold to define full TRAC; Green dashed lines (C): -15 Centiloid threshold to define “no TRAC”. Intraclass correlation coefficient and Pearson’s correlation are provided.

Among the 41 patients with paired PET (Table 2), the measured difference between scans (ΔCentiloids) also showed high agreement across pipelines (ICC= 0.916 [0.864, 0.951], all pairwise ICCs ≥ 0.89, Figure 1C). Using linear mixed-effect models, estimates of Centiloid change were consistent across pipelines, ranging from -42.7 to -43.6 Centiloids/year (see Figure 2A). At the individual level, 40/41(98%) individuals showed significant TRAC, with a decline exceeding 15 Centiloids with all 3 pipelines (Supplementary Figure 1A). One lecanemab-treated participant showed no TRAC, i.e., ΔCentiloids within the test-retest variability (between -5.9 and +8.3 Centiloids depending on pipelines; Supplementary Figure 1B).

**Table 2.** Characteristics of the participants with paired amyloid PET scans.

|  | **Whole sample**<br>**(N=41)** | **Donanemab**<br>**subgroup (N=15)** | **Lecanemab**<br>**subgroup (N=26)** | **P-value\*** |
| --- | --- | --- | --- | --- |
| **Age, years** | 73.5 ± 6.4 | 74.9 ± 6.0 | 72.7 ± 6.6 | 0.28 |
| **Sex, n (%)** |  |  |  | 1 |
| Female | 24 (59%) | 9 (60%) | 15 (58%) |  |
| Male | 17 (41%) | 6 (40%) | 11 (42%) |  |
| **Education, years** | 17.4 ± 2.2 | 16.4 ± 2.5 | 18.1 ± 1.8 | **0.034** |
| **APOE ε4 status, n (%)** |  |  |  | 1 |
| Carrier | 21 (51%) | 8 (53%) | 13 (50%) |  |
| Non-carrier | 20 (49%) | 7 (47%) | 13 (50%) |  |
| **Diagnosis, n (%)** |  |  |  | 0.45 |
| MCI | 32 (78%) | 13 (87%) | 19 (73%) |  |
| Mild dementia | 9 (22%) | 2 (13%) | 7 (27%) |  |
| **CDR-SB** | 2.8 ± 1.4 | 2.9 ± 1.2 | 2.8 ± 1.6 | 0.78 |
| **MMSE** | 25.4 ± 3.0 | 25.7 ± 2.4 | 25.2 ± 3.2 | 0.55 |
| **Pre-ATT Centiloids** |  |  |  |  |
| MIMneuro | 78 ± 36 | 83 ± 40 | 75 ± 35 | 0.54 |
| rPOP | 76 ± 37 | 76 ± 36 | 77 ± 38 | 0.94 |
| CapAIBL | 85 ± 39 | 85 ± 39 | 85 ± 39 | 0.97 |
| **ARIA-E** |  |  |  | 0.73 |
| Any ARIA-E | 12 (29%) | 5 (33%) | 7 (27%) |  |
| No ARIA-E reported | 29 (71%) | 10 (67%) | 19 (73%) |  |
| **ARIA-H** |  |  |  | 0.72 |
| Any ARIA-H | 10 (24%) | 3 (20%) | 7 (27%) |  |
| No ARIA-H reported | 31 (76%) | 12 (80%) | 19 (73%) |  |
| **Treated duration, months** | 16.7 ± 4.6 | 14.0 ± 4.1 | 18.2 ± 4.2 | **0.003** |

Demographic and clinical characteristics describe baseline (pre-ATT) state.
Missing values: n=1 for education, n=3 for CDR-SB.
P values: lecanemab versus donanemab subgroup comparison using two sample t-test (continuous variables) or Fisher exact test (categorical variables).

**Figure 2.**
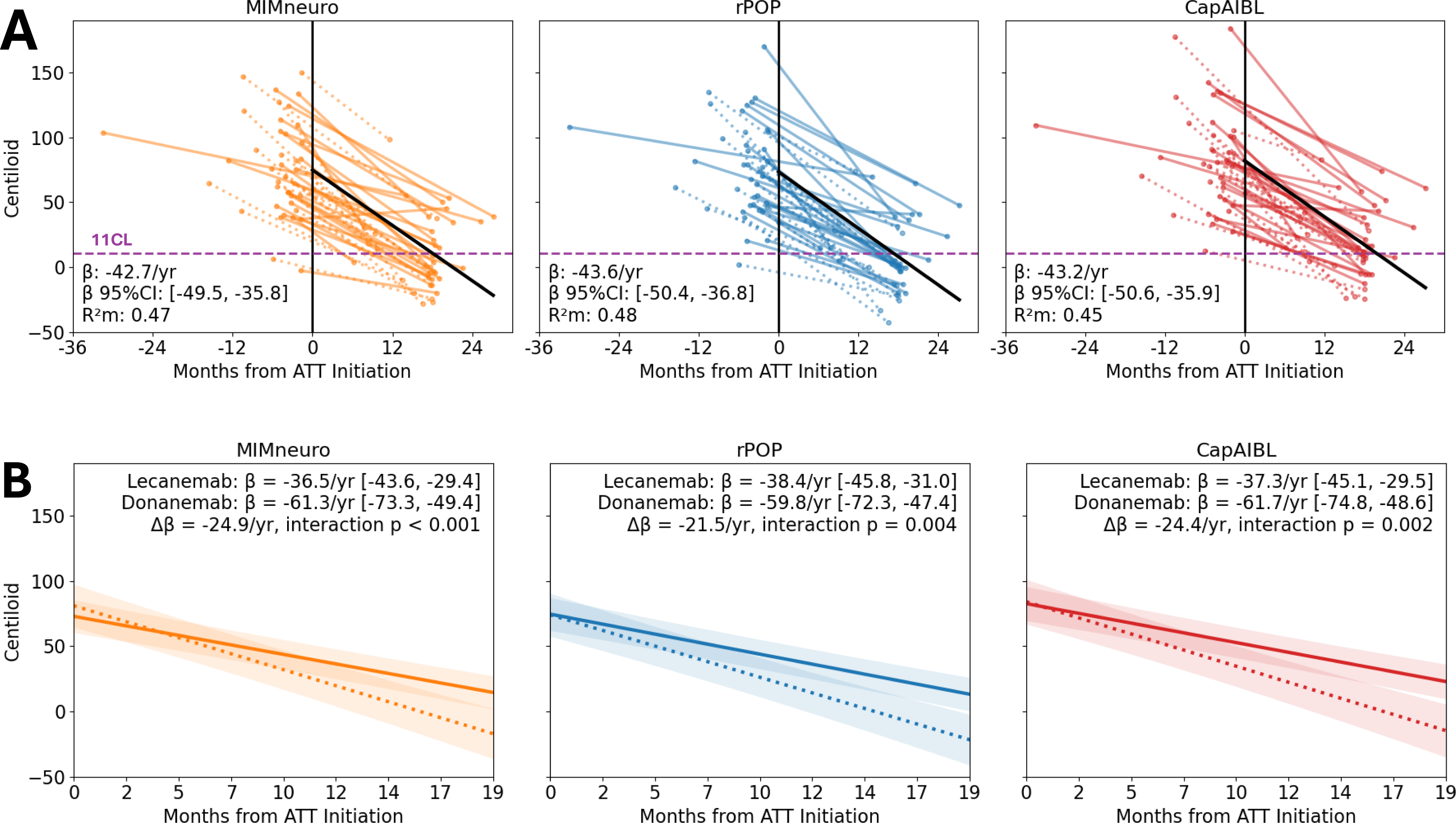
Rates of amyloid clearance estimated in patients with paired scans. A) Whole sample (n=41). Thin lines: raw trajectories (solid lines: lecanemab-treated individuals; dotted lines: donanemab-treated individuals), thick black lines: model estimates for amyloid clearance rates (β). R^2^m: Marginal R², variance in Centiloids explained by the fixed effect of time. Purple dashed line: 11 Centiloid threshold. B) Outcome of the linear mixed-effect models that included an interaction term between time and ATT. Solid lines: lecanemab, dotted lines: donanemab.

Subgroups treated with lecanemab (n=26) versus donanemab (n=15) were comparable in terms of age, sex, *APOE* genotype, clinical severity, and pre-ATT Centiloid values; patients treated with donanemab had slightly fewer years of education and underwent post-ATT PET ∼4 months earlier than lecanemab-treated participants (Table 2). Across pipelines, Centiloid decline was significantly greater in the donanemab-treated group (between -59.8 and -61.7 Centiloids/year), compared to lecanemab (−36.5 to -38.4 Centiloids/year, all time*treatment interactions: p<0.005, see Figure 2B).

### Centiloid-versus Visual Read-based classification of post-ATT scans

Among the 54 post-ATT scans (all 18F-florbetapir), approximately half fulfilled the quantitative criterion for full TRAC (<11 Centiloids): between 24 (44%) and 31 (57%) depending on pipeline (Figure 3A, Fleiss’ kappa = 0.68 [0.52 to 0.85]). Full agreement between pipelines was observed for 41 (76%) scans: n=21 scans that were ≥11 Centiloids with all 3 pipelines and n=20 that were <11 Centiloids with all 3 pipelines.

**Figure 3.**
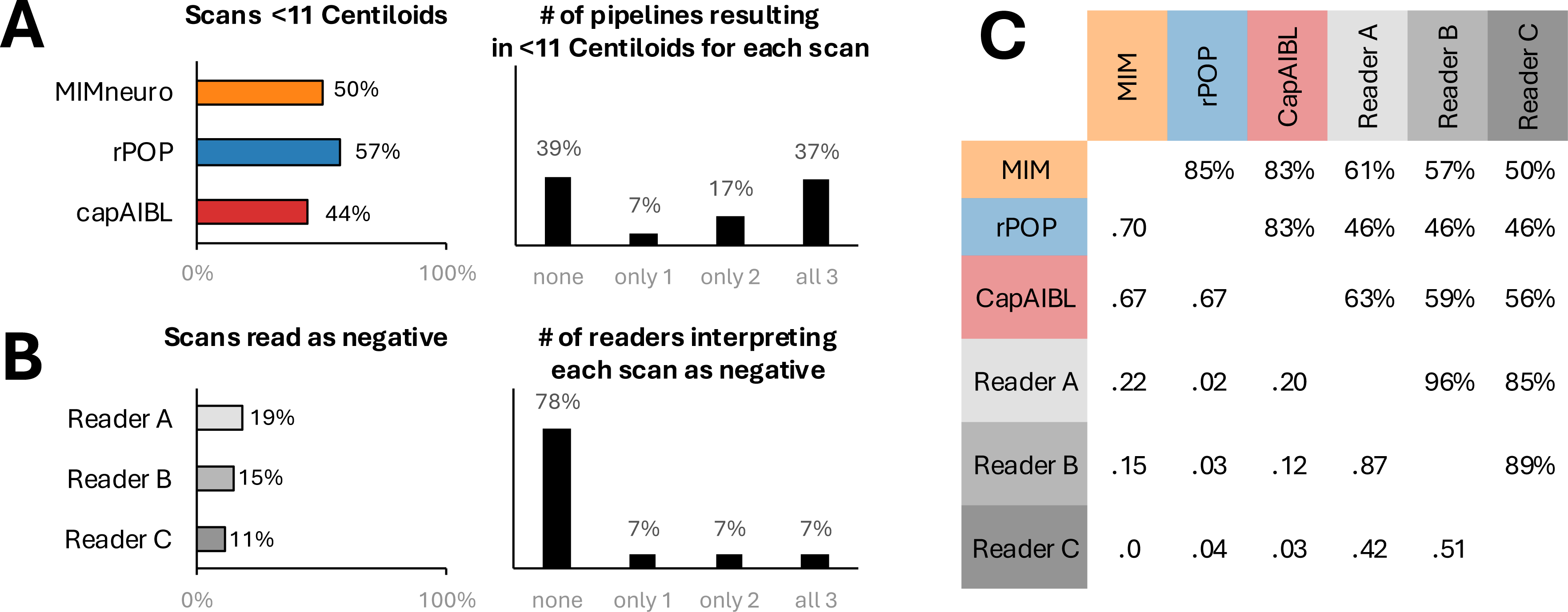
Full Treatment-Related Amyloid Clearance (TRAC) in scans acquired after ATT initiation (n=54) A) Full TRAC based on quantitation. B) Full TRAC based on visual reads; right panel percentages sum up to 99 due to rounding. C) Pairwise agreement between the 6 binary classifications (kappa below diagonal, percent agreement above).

In contrast, few post-ATT scans were visually read as negative: between 6 (11%) and 10 (19%) across raters (Figure 3B, Fleiss’ kappa = 0.61 [0.38 to 0.87]). Full inter-rater agreement was observed for 46 (85%) scans: 42 were unanimously read as positive and 4 as negative.

Pairwise agreement across all 6 approaches (3 pipelines and 3 readers) is illustrated in Figure 3C and detailed in Supplementary Table 1; specific examples of extreme discordance between Centiloid and visual reads are illustrated in Figure 4. Among the 20 scans that were <11 Centiloids with all 3 pipelines, 18 (90%) were read as positive by at least one reader and 13 (65%) were read as positive by all 3 readers. Readers noted frequent residual uptake in these scans (Figure 4A-B and Supplementary Figure 2), most commonly in the occipital/posterior cortex as noted in 13 (65%) scans by at least one reader. In contrast, 2 out of the 4 scans that were unanimously read as negative remained ≥11 Centiloids with all 3 pipelines (Figure 4C, Supplementary Table 1).

**Figure 4.**
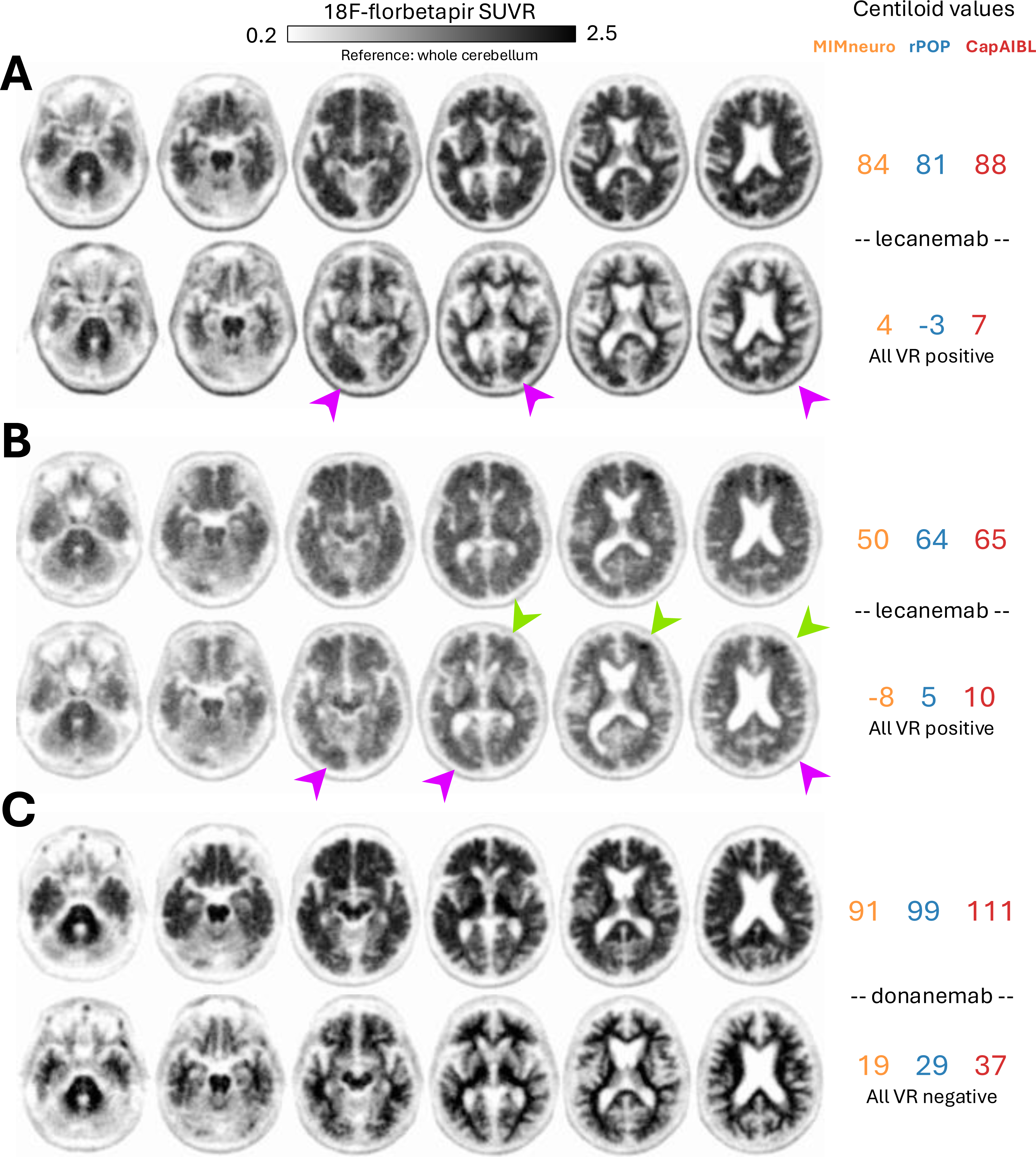
Examples of post-ATT amyloid PET with discordance between Centiloid quantitation and visual reads. Each panel shows pre-ATT (top) and post-ATT (bottom) PET for one participant. All scans were acquired using 18F-florbetapir on the same Biograph Vision scanner. VR: Visual Read. A-B: individuals with post-ATT quantitation <11 Centiloids across pipelines but positive by all 3 readers due to residual posterior (purple arrows) or frontal (green arrows) signal. C: individual whose post-ATT quantitation remained elevated (all ≥19 Centiloids) but was read as negative by all 3 readers.

### Complementary analyses in IDEAS

In the IDEAS dataset of 2,265 18F-florbetapir scans quantified below 11 Centiloids using rPOP, 365 (16.1%) were interpreted as positive by local readers.

## Discussion

Using data from our infusion clinic, we characterized the reliability of amyloid PET interpretation after ATT initiation. Centiloid values from 3 independent pipelines showed high agreement and yielded consistent quantitative outcomes. However, an unexpectedly high proportion of scans below a stringent 11 Centiloid threshold were interpreted as visually positive by expert readers. Discordance between visual reads and Centiloid quantitation is not uncommon in ATT-free individuals(*19–21*), but the magnitude of the discordance we observed is substantially higher than previous reports. Our complementary analyses demonstrated that the proportion of post-ATT scans <11 Centiloids that were visually read as positive (65%–90%, depending on the reader) greatly exceeded the expected rates in ATT-naive scans (16%).

Residual occipital binding appears to be a major contributor to this discordance: it was noted by at least one reader in 13/20 scans that were quantified <11 Centiloids by all 3 pipelines. The occipital lobe is not included in the masks traditionally used to measure amyloid PET signal (neither the default Centiloid mask(*6*), nor the Clark atlas(*22*) implemented in MIMneuro), because, at the group level, amyloid PET signal is usually lower in the occipital lobe than in other cortical areas(*6*,*23*). Previous publications have highlighted this issue in the context of visual/centiloid discordance (*20*,*24*,*25*), especially as recent investigations of large samples of ATT-naive individuals have identified a subgroup with predominant occipital binding (*26*,*27*) for whom typical Centiloid calculation underestimates amyloid burden. Moreover, the interpretation of occipital binding is variable across visual read methodologies, as the labels for 18F-florbetaben and 18F-flutemetamol exclude the occipital lobe from their regions of interest. Some of the post-ATT scans shown in the current study would likely not fulfill criteria for amyloid PET positivity if scans were read according to these methods rather than 18F-florbetapir’s approach. More generally, because Centiloid values average signal across a large cortical mask, focal residual binding can coexist with a globally low Centiloid value; regional quantitation, including of the occipital cortex, may therefore be a useful adjunct when interpreting post-ATT scans.

Residual occipital PET signal has been documented in an aducanumab-treated case who already showed a clear pre-ATT occipital-predominant pattern; autopsy confirmed that the highest density of residual plaques was observed in the occipital cortex(*28*). However, some patients in our study showed a different pattern, with globally positive pre-ATT scans and post-ATT scans showing focal occipital binding (Figure 4A), suggesting slower clearance in the occipital cortex compared to other areas. The biological interpretation of occipital signal in ATT-naive PET remains debated, but converging evidence suggests it is associated with the presence of cerebral amyloid angiopathy (CAA) (*27*,*29–31*). This hypothesis is particularly relevant in the context of ATT, not only because CAA is a risk factor for amyloid-related imaging abnormalities, but also because recent autopsy studies of aducanumab-treated individuals suggest that ATTs do not reduce, and may actually enhance amyloid deposits in microvessels(*32–34*). Post-ATT occipital PET binding may thus reflect residual or enhanced vascular amyloid deposits. Future research including PET and postmortem assessment of ATT-treated individuals will be needed to advance our understanding of this amyloid PET pattern.

Additional factors may contribute to the visual/Centiloid discordance we observed. ATTs are associated with accelerated brain volume loss and ventricular enlargement, thought to partly reflect removal of amyloid and associated inflammatory changes rather than neurodegeneration (“pseudo-atrophy”)(*35*). Because of partial-volume effects, such changes may both affect the gray/white contrast on which visual reads rely and impact the cortical quantitative measures underlying Centiloid calculation. Changes in cerebral blood flow could also affect tracer delivery and kinetics.

Centiloid-based outcomes were consistent across pipelines, including rates of Centiloid change and identification of the patient with no TRAC. Agreement on the status of post-ATT scans (full TRAC or not) was lower (Fleiss’ kappa=0.68), with pipelines disagreeing for 13 (24%) scans, reflecting the sensitivity of any fixed threshold to small inter-pipeline differences when post-ATT values cluster near the cutoff. Moreover, clearance estimates replicated values reported in phase 3 trials of lecanemab (between 36 and 38 Centiloids/year across pipelines versus 37 Centiloid/year in the trial(*2*)) and donanemab (between 60 and 62 Centiloids/year versus 58 Centiloids/year in the trial (*1*)). These results indicate that widely used Centiloid tools, including the FDA-cleared MIMneuro, yield clearance estimates comparable to those reported in clinical trials, supporting the robustness of Centiloid implementation.

The present work has limitations. First, this single-site study mainly used 18F-florbetapir (exclusively for post-ATT scans), which may limit generalizability. Our sample was relatively small, limiting our ability to more comprehensively characterize post-ATT discordant cases. Second, visual reads were not blinded to treatment status: the 3 expert readers knew scans were post-ATT, although they had no other information. Finally, our study included 3 PET-only quantitative pipelines, so it is unclear whether our conclusions could be extrapolated to other software or methods, especially pipelines that include structural imaging (CT or MRI) to improve anatomical positioning of the regions of interest.

In conclusion, we identified an unexpectedly high level of discordance between visual reads and Centiloid quantitation of amyloid PET scans after ATT, indicating that a low post-ATT Centiloid value might not, by itself, be equated with a negative scan. Our findings support integrating visual assessment, including attention to residual regional (particularly occipital) binding, with quantitation when reporting post-ATT scans. Ongoing efforts to collect PET scans from real-world clinical settings through large-scale repositories (*36*) will help answer questions that were not addressed in clinical trials and improve our use of amyloid PET as a treatment monitoring tool. The clinical relevance of the present findings remains to be established as it is unclear whether and how different degrees of TRAC should guide the optimal management of ATT-treated patients.

## Data Availability

All data produced in the present study are available upon reasonable request to the authors

## Acknowledgments

We are grateful to all patients and study partners.

## Key points

### QUESTION

Do Centiloid quantitation and visual reads agree when determining whether amyloid PET has become negative after amyloid-targeting therapy?

### PERTINENT FINDINGS

In patients treated with lecanemab or donanemab, Centiloid values and clearance rates were highly consistent across 3 pipelines. However, 13 of 20 post-treatment scans below 11 Centiloids with all pipelines were read as positive by all 3 readers, often because of residual occipital binding.

### IMPLICATIONS FOR PATIENT CARE

A low post-treatment Centiloid value should not by itself be equated with a negative scan; integrating visual reads, including assessment of residual regional binding, with quantitation is advisable when assessing treatment-related amyloid clearance.

