## supplements for "When Is Amyloid Really Cleared? Substantial Discordance Between Centiloid Quantitation and Visual Reads of Amyloid PET after Amyloid-Targeting Therapy"

### Supplementary Table 1. Individual scan information for the binary classification of the 54 post-ATT scans.

Each row represents 1 post-ATT scan; green cells indicate full TRAC (<11 Centiloids or negative read)

| MIMneuro | rPOP | CapAIBL | Reader A | Reader B | Reader C |
|---|---|---|---|---|---|
| 98 | 74 | 83 | positive | positive | positive |
| 38 | 26 | 28 | positive | positive | positive |
| 35 | 24 | 31 | positive | positive | positive |
| 39 | 39 | 59 | positive | positive | positive |
| 28 | 21 | 30 | positive | positive | positive |
| 54 | 36 | 30 | positive | positive | positive |
| 45 | 41 | 43 | positive | positive | positive |
| 15 | 16 | 30 | positive | positive | positive |
| 18 | 18 | 18 | positive | positive | positive |
| 30 | 15 | 23 | positive | positive | positive |
| 55 | 70 | 62 | positive | positive | positive |
| 53 | 64 | 74 | positive | positive | positive |
| 35 | 41 | 52 | positive | positive | positive |
| 39 | 48 | 61 | positive | positive | positive |
| 46 | 34 | 40 | positive | positive | positive |
| 17 | 14 | 27 | positive | positive | positive |
| 50 | 36 | 47 | positive | positive | positive |
| 42 | 34 | 42 | positive | positive | positive |
| 26 | 25 | 25 | positive | positive | positive |
| 19 | 29 | 37 | negative | negative | negative |
| 28 | 32 | 38 | negative | negative | negative |
| 17 | -3 | 33 | positive | positive | positive |
| 19 | 10 | 17 | positive | positive | positive |
| 15 | 0 | 16 | positive | positive | negative |
| 4 | 15 | 15 | negative | negative | positive |
| -10 | 1 | 12 | positive | positive | positive |
| 21 | 2 | 10 | positive | positive | positive |
| -5 | 0 | 16 | positive | positive | positive |
| 1 | 3 | 15 | positive | positive | positive |
| 9 | 9 | 21 | positive | positive | positive |
| 14 | -6 | 1 | positive | positive | positive |
| 0 | 5 | 12 | positive | positive | positive |
| 15 | 6 | 5 | positive | positive | positive |
| 1 | 16 | 8 | negative | positive | positive |
| -14 | -21 | 9 | positive | positive | positive |
| -10 | 0 | 2 | positive | positive | positive |
| -12 | -23 | -4 | positive | positive | positive |
| 4 | -3 | 7 | positive | positive | positive |
| -20 | -13 | -6 | positive | positive | positive |
| -6 | -27 | -14 | positive | positive | positive |
| -8 | 5 | 10 | positive | positive | positive |
| -1 | 6 | 7 | positive | positive | positive |
| -6 | -13 | -5 | positive | positive | positive |
| -3 | 0 | -5 | positive | positive | positive |
| 0 | -14 | 4 | positive | positive | positive |
| -27 | -26 | -25 | positive | positive | positive |
| -24 | -22 | -17 | positive | positive | positive |
| -14 | -20 | -13 | positive | positive | negative |
| -5 | 2 | 6 | negative | positive | positive |
| -13 | -16 | -12 | negative | negative | positive |
| -28 | -43 | -22 | negative | negative | positive |
| 9 | 0 | 7 | negative | negative | positive |
| -20 | -33 | -24 | negative | negative | negative |
| -9 | -20 | -5 | negative | negative | negative |

**Supplementary Figure 1. Individual patient with no Treatment Related Amyloid Clearance**.
A. Histogram of the change in Centiloids calculated for individuals with paired scans (n=41). The green dashed line indicates the 15 Centiloid threshold to operationalize "no TRAC" (change within test-retest variability). Across all three pipelines, 1 individual does not show evidence for TRAC.
B. Similar to Figure 2A; patient with no TRAC is indicated in black.
C. 18F-florbetapir scans for that individual. Both scans were acquired on the same Biograph Vision scanner with identical reconstruction parameters. Baseline scan was acquired 31-41 min post injection; follow-up scan was acquired 30-40 min post injection.

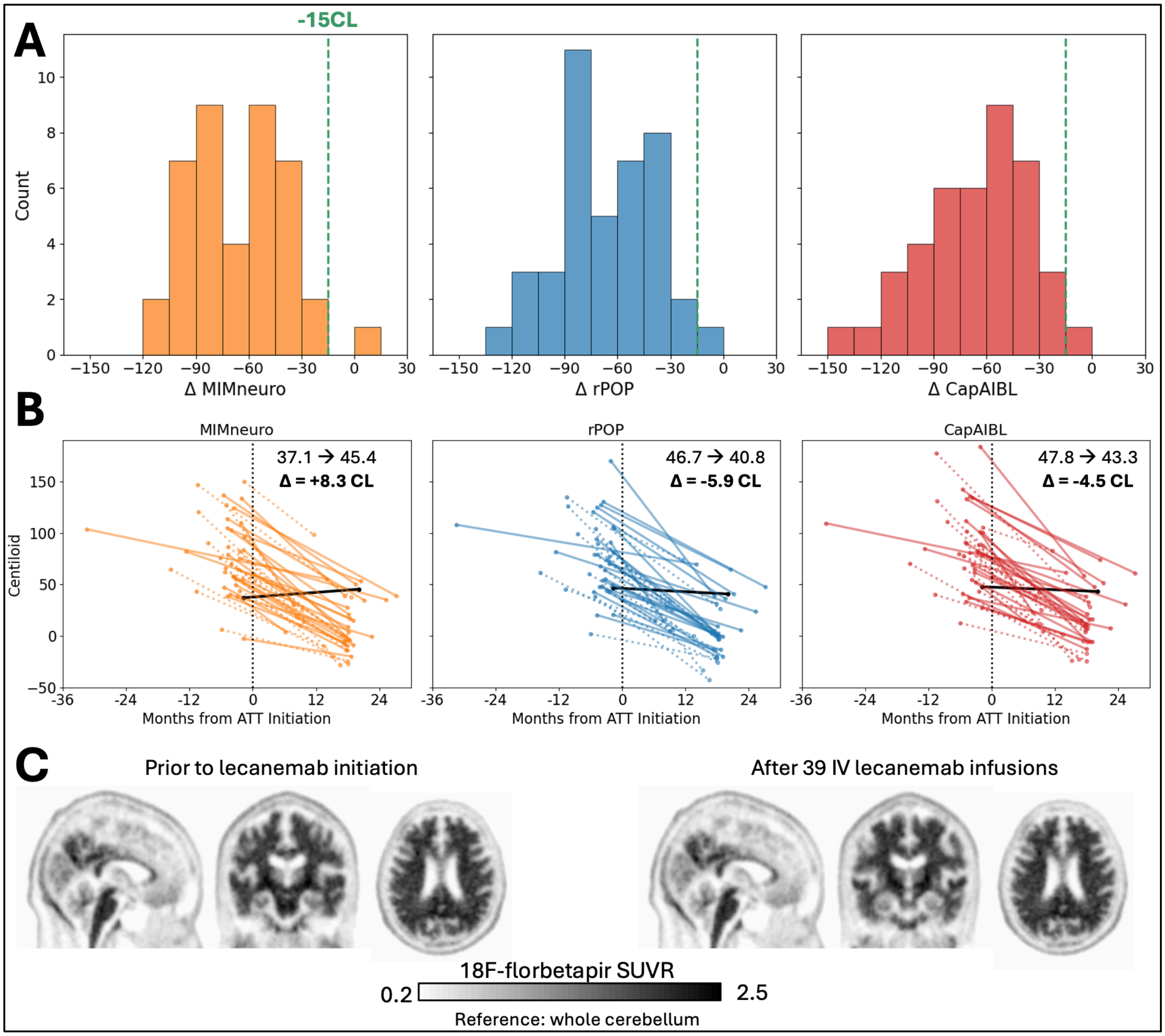

**Supplementary Figure 2. Five additional examples of post-ATT 18F-florbetapir scans that were read as positive by all 3 readers but had low quantitation across pipelines (<11 Centiloids).**
Purple arrows indicate residual occipital/posterior binding; green arrows indicate frontal binding.

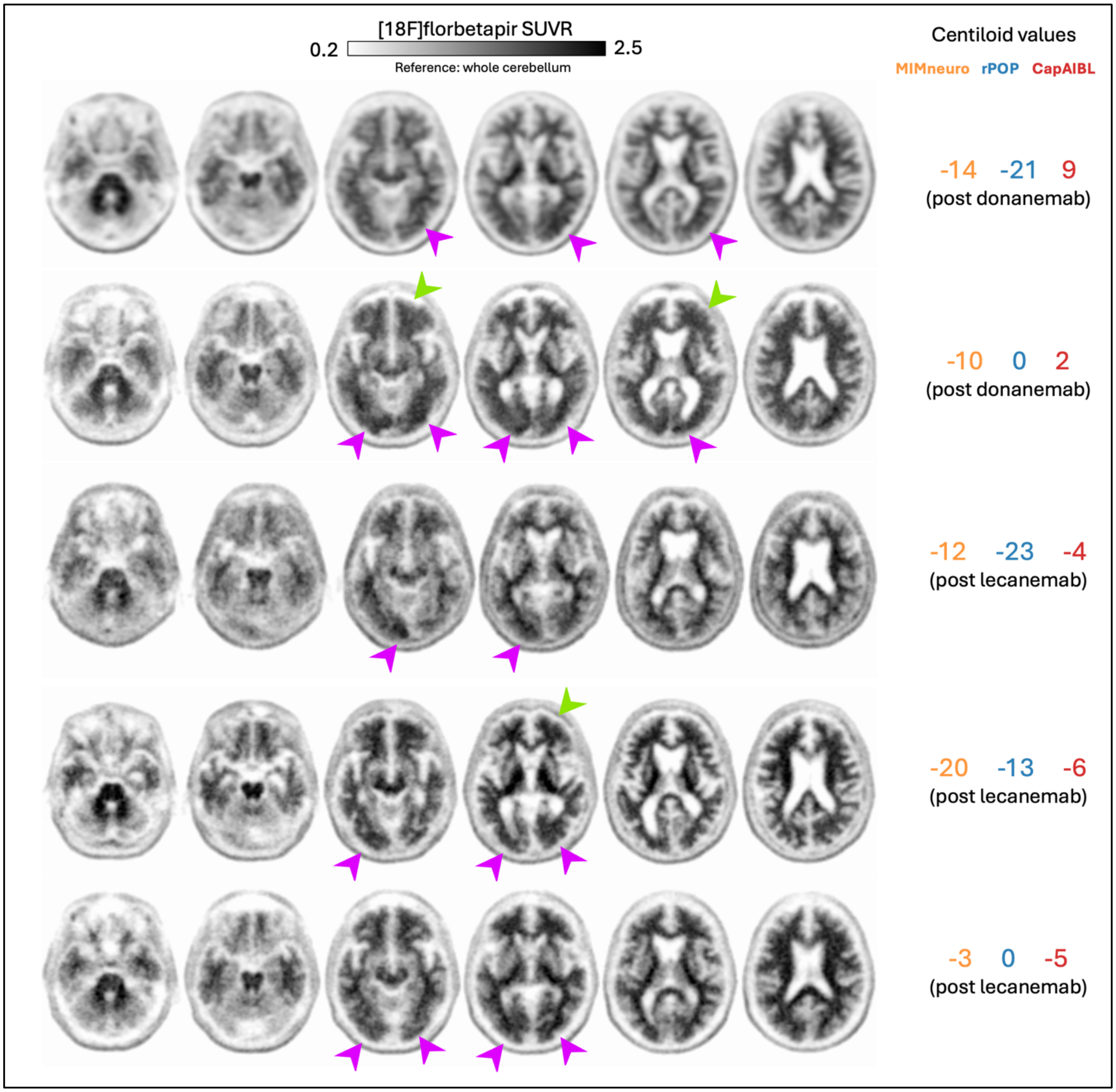
